# Data-driven modeling of trigeminal nerve shape reveals pain-associated morphology in trigeminal neuralgia

**DOI:** 10.64898/2026.11.11.26367487

**Authors:** Michael E. Xie, Haoyuan Liang, Xihang Wang, Vivek Yedavalli, Christopher Jackson, Michael Lim, Judy Huang, Chetan Bettegowda, Peirong Liu, Risheng Xu

## Abstract

Although the leading theory for the cause of trigeminal neuralgia (TN) is neurovascular compression, compression alone does not fully explain which nerves are painful. Recent work has demonstrated that pain in TN is associated with morphological changes of the trigeminal nerve, which may be a result of specific forms of compression or other disease processes. These changes have so far been quantified by hand-crafted geometric measurements, which can miss higher-order characteristics of the anatomy that are difficult to visualize. In this study, we use a cohort of 479 patients with TN who underwent microvascular decompression and had high-resolution pre-operative MRI scans to determine whether data-driven 3D shape features of the trigeminal nerve capture pain-associated morphology better than predefined metrics. We find that a predictive model using our shape features can distinguish nerves implicated in pain from those without pain in a held-out test cohort with an area under the receiver operating characteristic curve (AUROC) of 0.719, compared with 0.626 for a model using existing hand-crafted measurements (p=0.005). Our study identifies candidate nerve morphology biomarkers of TN pain through data-driven shape analysis and demonstrates that these higher-order features relate better to symptoms of TN than conventional shape metrics.

## Introduction

Trigeminal neuralgia (TN) is a debilitating facial pain disorder, characterized by sharp bouts of pain in the sensory distribution of the trigeminal nerve on one side of the face.[6,18] Despite the rarity of the disease, the severity of pain can be extraordinarily disruptive to patients’ wellbeing.[14]

The leading hypothesis for the cause of pain in TN is compression of the trigeminal nerve by nearby vasculature, and therefore, a common surgical treatment for TN is microvascular decompression (MVD). In this surgery, a pledget is placed between the trigeminal nerve and nearby vessels to prevent contact. Neurovascular contact alone, however, does not fully account for the pain experienced by patients, as 20–30% of patients experience recurrence of pain after MVD.[18] Additionally, studies have found that neurovascular contact is also prevalent on the asymptomatic side in unilateral TN patients.[13] A better understanding of other factors that relate to TN symptoms would be critical for understanding the underlying pathology and ultimately improve treatment.

Recently, the shape of the trigeminal nerve has been proposed as an important additional factor for characterizing pain in TN. Studies in support of this idea have noted that features such as torsion, curvature, cross-sectional area, and flatness of the nerve relate to whether the patient experienced pain in that nerve.[2,9,10] These papers have suggested that some factors affecting a nerve’s shape, like atrophy from demyelination and axonal loss or physical distortion of the nerve, may also drive pain.[7,9,13,17] Understanding the shape features that are the most relevant for pain, therefore, might reveal insights into how symptoms arise in TN.

Existing work analyzing trigeminal nerve shape has focused on anatomical features that were hand-crafted by clinicians. Such features can summarize specific characteristics of the nerve shape but may miss higher-order features, such as spatially distributed or covarying changes, that could also relate to pain. New methods to automatically segment the trigeminal nerve from pre-operative MRI data have made quantitative analysis of large patient cohorts much more efficient and realistic.[8,16] Using these tools, we sought to use a data-driven approach to discover three-dimensional (3D) shape features of the trigeminal nerve and see if such features could more accurately separate symptomatic from asymptomatic nerves than existing metrics.

## Methods

### Patient cohort

Trigeminal neuralgia patients who underwent primary MVD at our institution between January 2014 and December 2019 were identified (IRB00338945). Patients were included in this study if they had pre-operative high-resolution Constructive Interference in Steady State (CISS) or Fast Imaging Employing Steady-state Acquisition (FIESTA) brain MRI sequences available.[1] Patients were then randomly split into a training cohort (80%) for development of a shape model and a testing cohort (20%) for evaluation of the shape model.

### MRI pre-processing and nerve segmentation

MRI data for all patients were pre-processed as in Wang et al.[16] The centroid of each of the patients’ nerves was identified by a trained researcher and verified by an attending neurosurgeon. A 30 mm x 30 mm x 30 mm volume around each centroid was then cropped out and voxels were rescaled to an isotropic size of 0.47 mm in each dimension. Intensity values of voxels within each crop were then Z-scored. Within each cropped volume, the nerve was segmented automatically using an nnU-Net model that was previously trained to segment trigeminal nerves on a distinct dataset.[16]

To later compare the shapes of the nerves, we aligned each nerve segmentation to a common orientation. First, left-side nerves were mirrored across the sagittal plane, so all nerves had the same medial to lateral axis direction. The principal axes of each nerve were then computed from the coordinates of the voxels of its segmentation, and nerve segmentations were rotated so that the principal axes were aligned across all nerves. The centroids of all segmentations were also aligned to the same location.

### Calculating conventional shape metrics

Using our trigeminal nerve segmentations, we first calculated a set of metrics proposed by Ishiwada et al. for each nerve (“conventional shape metrics”): centerline length, curvature, torsion, cross-sectional area, and flattening ratio.[10] We did not include the long-axis angle since our nerves were reoriented during alignment. The centerline of each nerve was extracted by a volumetric skeletonization algorithm.[12] The centerline length was then calculated as the sum of the distances between consecutive points along this centerline. Curvature and torsion were also calculated at each point of the centerline and the median value was used to represent the nerve, as in Ishiwada et al.[10] At each point of the centerline, cross-section was defined as the intersection of the nerve with the plane perpendicular to the centerline. For each nerve, the median area of the cross-sections along the nerve and the median flattening ratio (1–[short axis length]/[long axis length]) of the cross-sections were also recorded.

### Trigeminal nerve shape model

To build a statistical model of the nerve shape, each segmentation was converted to a signed distance field (SDF), where each voxel stores its distance to the nerve surface, with negative values indicating points inside the nerve and positive values indicating points outside. We performed principal component analysis (PCA) on the SDFs of the nerves in the training cohort. We kept the minimum number of principal components (PCs) needed to explain at least 85% of the variance in nerve shape in the training cohort. The scores of each nerve on these PCs were used as its shape features.

Nerves in the testing cohort were then projected onto these PCs to evaluate their relationship to the conventional shape metrics and disease characteristics.

### Model for distinguishing symptomatic and asymptomatic nerves

We used a logistic regression model both with and without L2 regularization to predict whether each nerve was symptomatic or asymptomatic from its 10 shape features. The unregularized model was trained on the training cohort to assess the effects of each shape feature on symptomatic identity in a multivariate model. The regularized model was trained on the training cohort and evaluated on the testing cohort to assess generalizable prediction performance. Performance was measured by the area under the receiver operating characteristic curve (AUROC). To compare our shape features with the conventional shape metrics, a second model was trained in the same way on the five conventional metrics. We also fit a model including PC scores and conventional shape metrics all together as predictors and a model using total nerve volume alone as a predictor. For all models, features were standardized using the mean and standard deviation of the training cohort. We estimated 95% confidence intervals for the AUROC from 2,000 bootstrap resamples of the patients in the testing cohort. The difference in AUROC between the models was tested with a paired bootstrap. We also ran a permutation test (500 permutations) to test whether each model performed better than chance. In each permutation, the labels of the two nerves were swapped in about half of the patients, chosen at random, and the model was trained and tested again.

### Statistical analysis

To compare metrics between two paired populations, two-sided Wilcoxon signed-rank tests were performed. Logistic regression coefficients in the unregularized model were tested against zero with a Wald test using standard errors clustered by patient, since each patient contributed two nerves. All analyses were performed in Python 3.10.20 using NumPy 2.0.1, SciPy 1.15.2, scikit-image 0.25.2, scikit-learn 1.7.2, statsmodels 0.14.6, and pandas 2.3.3.

## Results

### Patient characteristics

A total of 479 patients underwent MVD during the study period and had pre-operative high-resolution CISS/FIESTA MRI imaging data available. The average age of patients was 56.3 ± 0.7 and 321 (67.0%) were female (**Table 1**). Of the patients, 192 (40.1%) had left-sided pain and the remainder had right-sided pain. No patients had bilateral TN pain. 404 (84.3%) patients experienced Type I pain and 74 (15.4%) experienced Type II pain. One patient experienced both types of pain. For our study, patients with the different types of pain were pooled together.

**Table 1:** Patient demographic information. Demographic information for 479 patients included in the study.

|  | **N=479** |
|---|---|
| Age at surgery, years |  |
| Mean ± s.e.m. | 56.3 ± 0.7 |
| Sex |  |
| Female | 321 (67.0%) |
| Male | 158 (33.0%) |
| Laterality of symptoms |  |
| Left | 192 (40.1%) |
| Right | 287 (59.9%) |
| TN type |  |
| Type 1 only | 404 (84.3%) |
| Type 2 only | 74 (15.4%) |
| Type 1 and 2 | 1 (0.2%) |

### Geometric measurements of trigeminal nerves

We first characterized the shape of the trigeminal nerve in our patients using the set of geometric measurements introduced by Ishiwada et al. [10] and compared the metrics between nerves on the symptomatic and asymptomatic sides (**Table 2**). The average cross-sectional area was smaller on the symptomatic side (6.353 mm^2^ vs. 6.775 mm^2^, p<0.001), and the average flattening ratio was larger on the symptomatic side (0.357 vs. 0.316, p<0.001). Nerves on the symptomatic and asymptomatic side were not significantly different in terms of centerline length, curvature, or torsion.

**Table 2:**
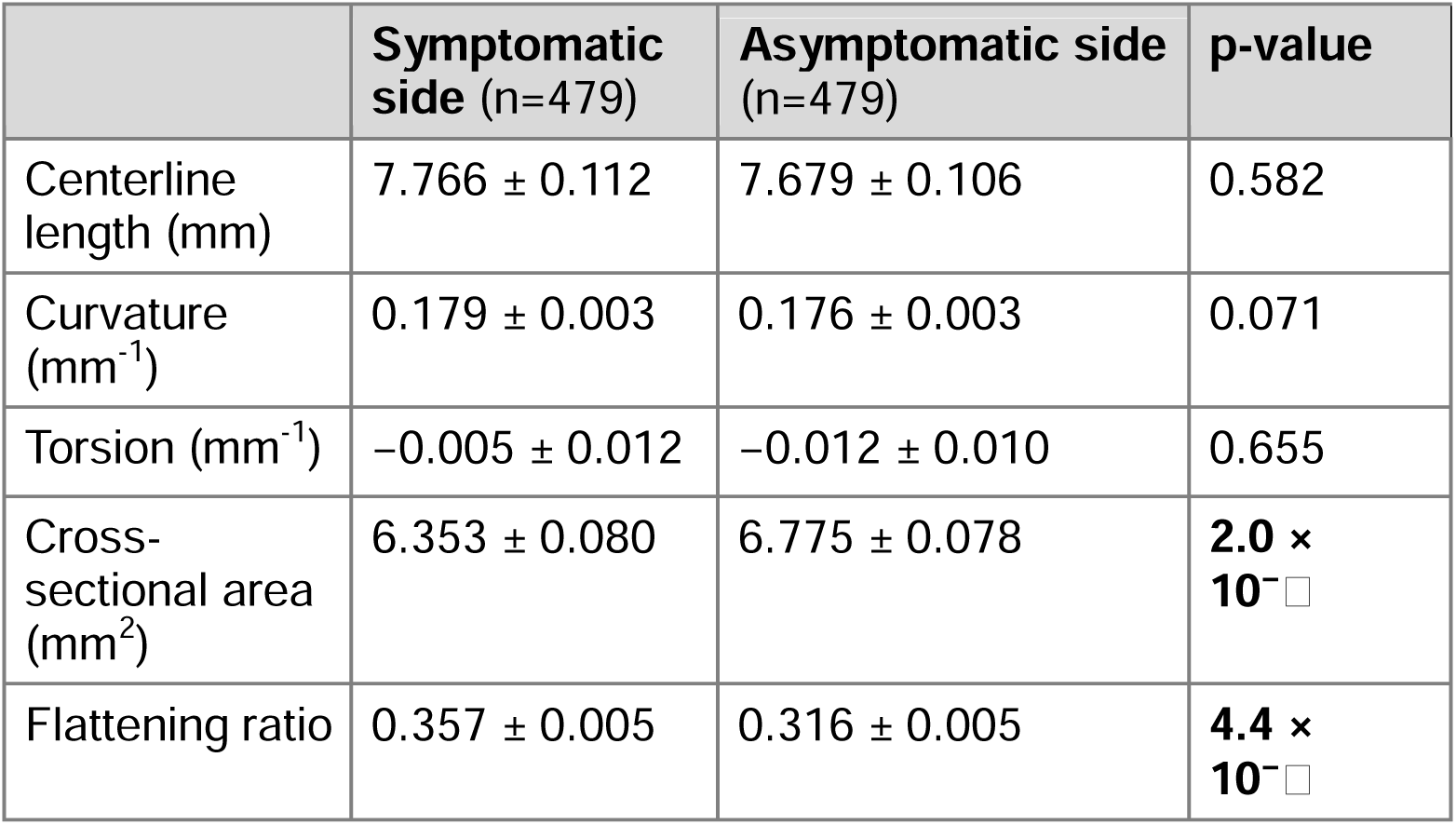
Morphometric comparison of symptomatic and asymptomatic nerves. Values are mean ± s.e.m. for 958 nerves from 479 patients. Symptomatic and asymptomatic nerves of the same patient were compared with Wilcoxon signed-rank tests (paired, two-sided).

|  | **Symptomatic side** (n=479) | **Asymptomatic side** (n=479) | **p-value** |
|---|---|---|---|
| Centerline length (mm) | 7.766 ± 0.112 | 7.679 ± 0.106 | 0.582 |
| Curvature (mm<sup>-1</sup>) | 0.179 ± 0.003 | 0.176 ± 0.003 | 0.071 |
| Torsion (mm<sup>-1</sup>) | −0.005 ± 0.012 | −0.012 ± 0.010 | 0.655 |
| Cross-sectional area (mm<sup>2</sup>) | 6.353 ± 0.080 | 6.775 ± 0.078 | **2.0 × 10⁻[illegible]** |
| Flattening ratio | 0.357 ± 0.005 | 0.316 ± 0.005 | **4.4 × 10⁻[illegible]** |

### Data-driven discovery of trigeminal nerve shape features

We next used PCA to find a set of features that capture the diversity of trigeminal nerve shapes across patients. The PCA was performed on the 766 nerves from 383 (80%) patients that made up the training data. For the analysis to be invariant to patient positioning in the scanner, all nerves were aligned along their principal axes. The longest principal axis represents the length of the nerve from the root entry point to the porus trigeminus. The median axis represents what we denote as the width of the nerve, and the shortest principal axis represents what we denote as the thickness of the nerve. We found that a set of 10 principal components (PCs) was the smallest set of components that captured at least 85% of the variance in shape across the trigeminal nerves in the training cohort (**Figure S1**). These 10 PCs were denoted as our trigeminal nerve shape features (**Figure 1A**).

**Figure 1:**
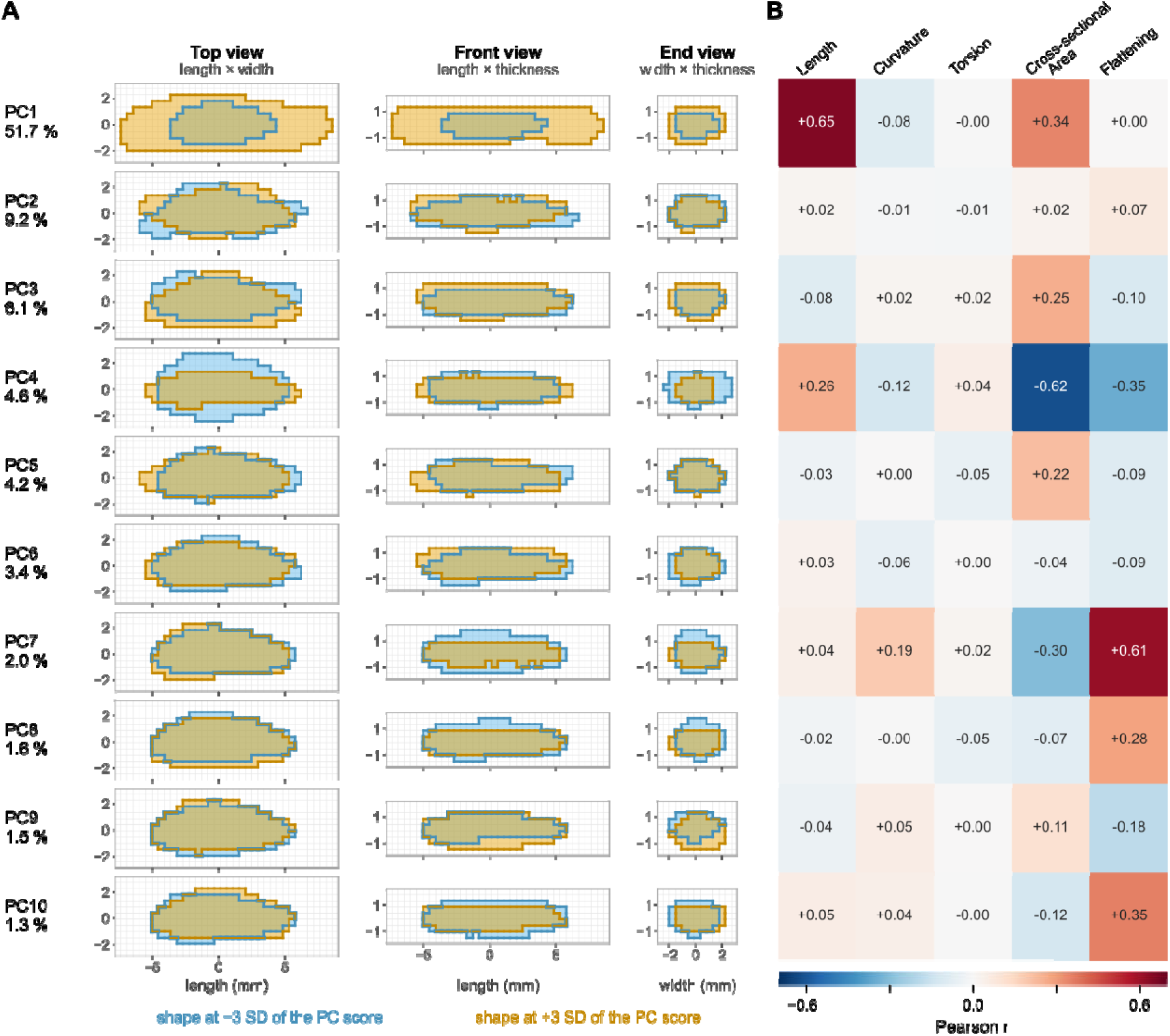
Statistical shape model (principal components of nerve segmentations). (A) Each of the 10 principal components (PCs) in the three views (defined in **Figure S2**), as the shape at −3 SD (blue) and at +3 SD (yellow) of its score. The percentage beside each PC is the proportion of population shape variance that PC explains. (B) Pearson correlation between each PC score and the five conventional shape metrics, calculated on the 766 nerves in the training data.

To understand what interpretable characteristics our shape features might represent, we correlated the PC scores of each nerve to its conventional shape metrics (**Figure 1B**). We found that several PCs shared information with interpretable geometric measures. For example, PC1 represented an overall size feature of the nerve, related to length and cross-sectional area. PC4 and PC7 were highly related to cross-sectional area and flattening ratio, and examination of the variations in shape shows that PC4 appears to represent a widening of the nerve, whereas PC7 relates more to a decrease in the thickness of the nerve.

### 3D shape features correlate with trigeminal nerve pain

Since we found that the PCs were correlated to conventional shape metrics that related to the nerve being on the symptomatic side, we sought to see if any of the PCs also were related to symptoms in the nerve. We assessed the relationship between PC scores and nerve symptoms in the held-out testing cohort of 96 patients (**Table 3**). The PC score differed significantly between trigeminal nerves in the symptomatic and asymptomatic side for PC1, PC7, and PC8. These PCs were ones that were associated with the overall nerve size and the thickness and flatness of the nerve.

**Table 3:**
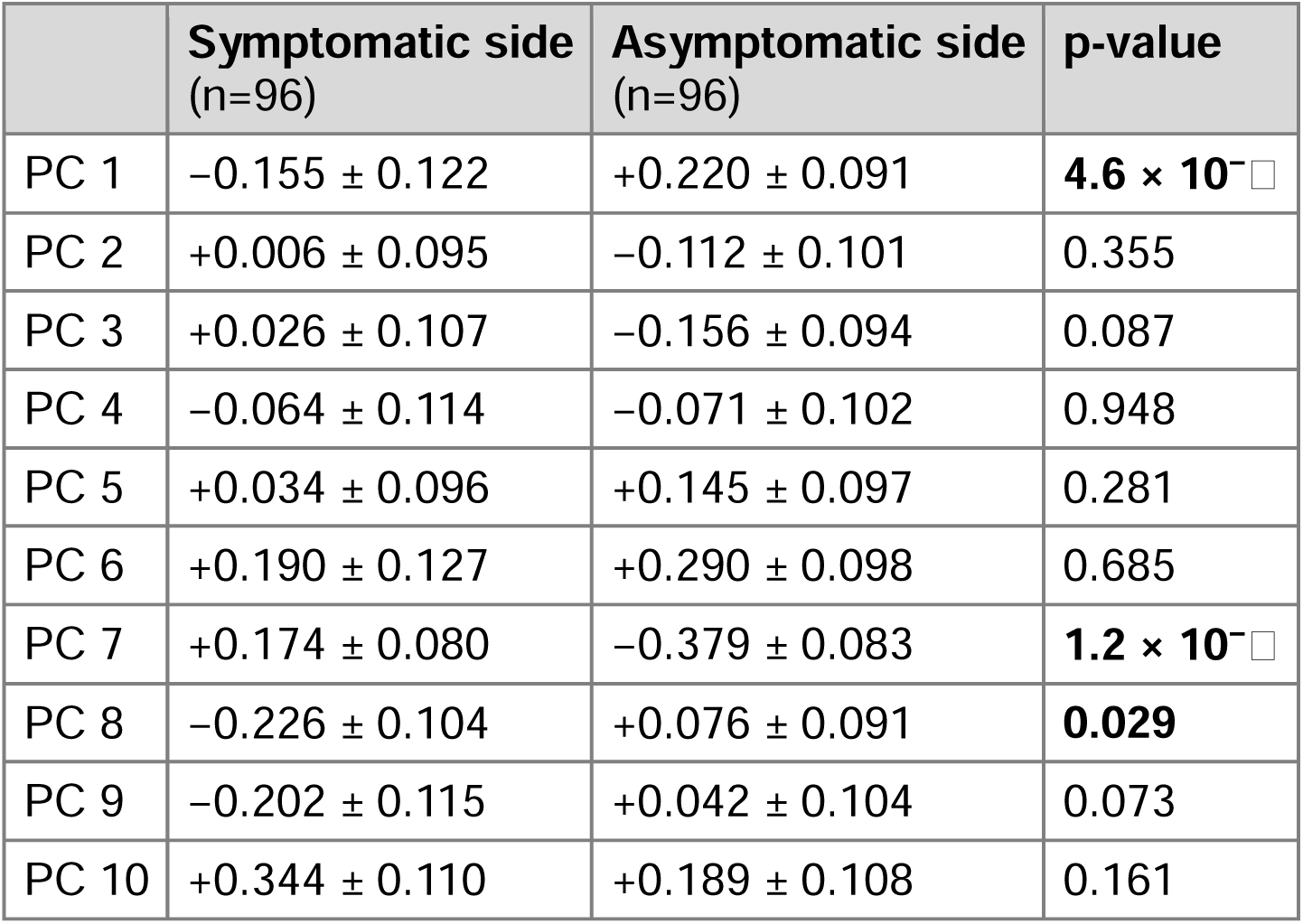
PC scores for nerves on the symptomatic side vs. asymptomatic side. Symptomatic and asymptomatic nerves of the same patient were compared with Wilcoxon signed-rank tests (paired, two-sided).

|  | **Symptomatic side**<br>(n=96) | **Asymptomatic side**<br>(n=96) | **p-value** |
|---|---|---|---|
| PC 1 | −0.155 ± 0.122 | +0.220 ± 0.091 | **4.6 × 10<sup>−[illegible]</sup>** |
| PC 2 | +0.006 ± 0.095 | −0.112 ± 0.101 | 0.355 |
| PC 3 | +0.026 ± 0.107 | −0.156 ± 0.094 | 0.087 |
| PC 4 | −0.064 ± 0.114 | −0.071 ± 0.102 | 0.948 |
| PC 5 | +0.034 ± 0.096 | +0.145 ± 0.097 | 0.281 |
| PC 6 | +0.190 ± 0.127 | +0.290 ± 0.098 | 0.685 |
| PC 7 | +0.174 ± 0.080 | −0.379 ± 0.083 | **1.2 × 10<sup>−[illegible]</sup>** |
| PC 8 | −0.226 ± 0.104 | +0.076 ± 0.091 | **0.029** |
| PC 9 | −0.202 ± 0.115 | +0.042 ± 0.104 | 0.073 |
| PC 10 | +0.344 ± 0.110 | +0.189 ± 0.108 | 0.161 |

### 3D shape features improve prediction of pain from trigeminal nerve shape

To understand how well our set of PCs together captured information associated with whether a nerve was associated with pain, we trained a logistic regression model on our training cohort (**Table 4**). In this multivariate analysis, PC1, PC3, PC7, and PC9 were statistically significantly related to pain in the nerve. To evaluate the generalizability of this model in predicting pain, we then tested the model on nerves from the patients in the testing cohort. On the held-out data, the model achieved an area under the receiver operating characteristic curve (AUROC) of 0.719, which was significantly greater than chance (**Figure 2**). We separately fit a logistic regression model to predict pain from the set of conventional shape metrics from Ishiwada et al.[10] using the training cohort, and that model achieved an AUROC of 0.626 on the test cohort. Compared to the Ishiwada-based model using the conventional metrics, our model was able to distinguish the nerves with pain and without pain in a significantly improved manner (AUC_PCs_– AUC_conventional_: 0.093, p=0.005). A combined model including both PC scores and conventional shape metrics as predictors also performed significantly better than the Ishiwada-based model (AUC_combined_–AUC_conventional_: 0.082, p=0.002) and performed similarly to the PC only model (AUC_combined_–AUC_PCs_: –0.011, p=0.53) (**Figure S3A**).

**Figure 2:**
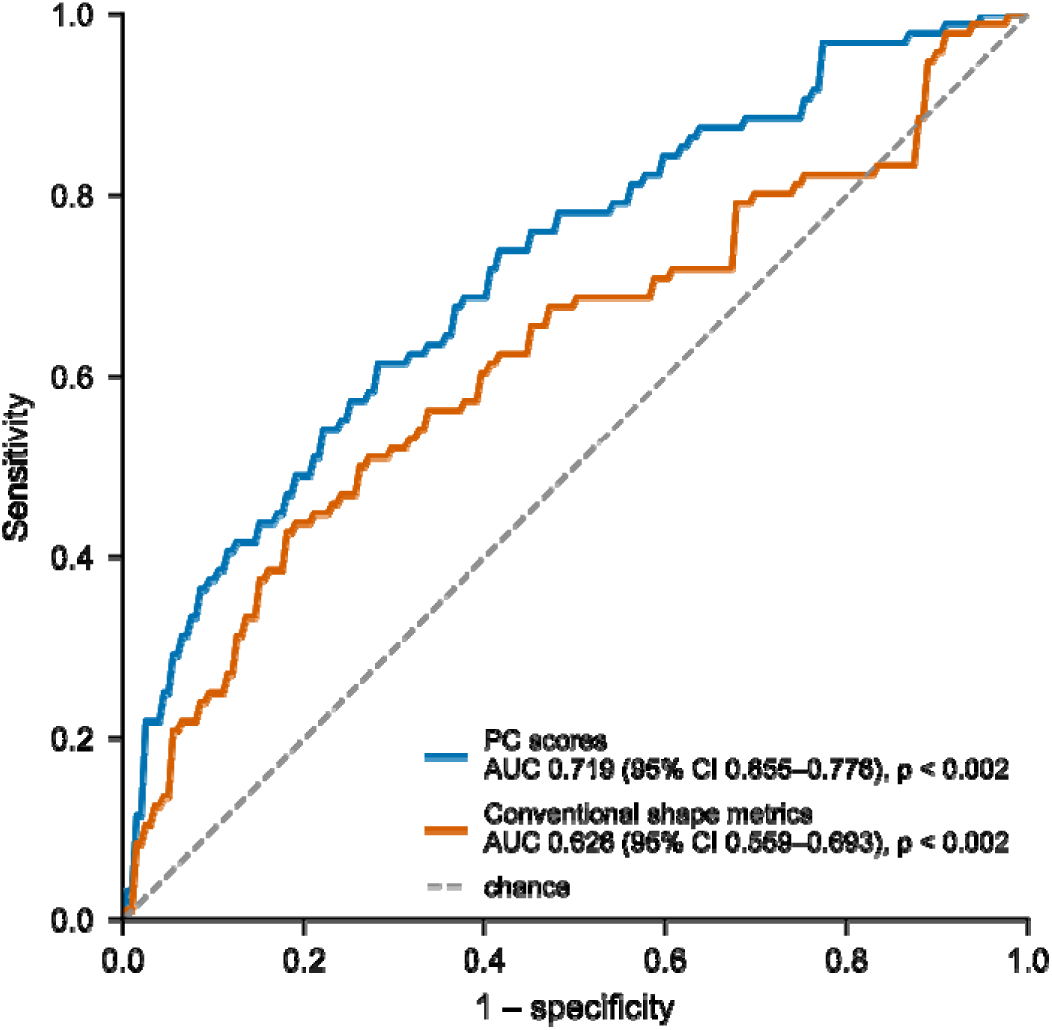
Predictive model for symptomatic nerve identity. Receiver-operating characteristic (ROC) curves comparing logistic regression models for predicting whether a test cohort nerve is on the symptomatic side using the PC scores (blue) or conventional shape metrics (orange). Hypothesis testing assessed whether AUCs were greater than 0.5 (chance).

**Table 4:** Multivariate relationship between 3D shape features and symptomatic nerve identity. A multivariate model was fit on the training cohort and coefficients for each predictor were tested against zero with a Wald test.

| **Predictor** | **OR (per 1 SD)** | **p-value** |
|---|---|---|
| PC1 | 0.892 | **0.025** |
| PC2 | 1.121 | 0.112 |
| PC3 | 0.844 | **0.017** |
| PC4 | 0.996 | 0.952 |
| PC5 | 0.912 | 0.183 |
| PC6 | 0.941 | 0.415 |
| PC7 | 1.621 | **1.23 × 10<sup>-10</sup>** |
| PC8 | 0.878 | 0.058 |
| PC9 | 0.852 | **0.031** |
| PC10 | 0.991 | 0.899 |

Since PC1 largely reflected nerve size, we also assessed whether nerve size alone could explain the predictive value of our PCs. A model using only total nerve volume achieved a significantly lower AUC than our model using the full set of PCs (AUC_volume_– AUC_PCs_: –0.126, p<0.001) (**Figure S3B**).

## Discussion

In this study, we leveraged a large cohort of TN patients to develop data-driven trigeminal nerve shape features and found that these new features were more strongly associated with pain in a nerve compared to previously hand-crafted geometric metrics. A benefit of our data-driven shape features is that they can capture nuanced shape features that are not easily observable or interpretable by humans. Additionally, empirical approaches are less biased by preconceptions of what features are likely to relate to pain. Therefore, our set of predictors can encode more information about trigeminal nerve shape than previously proposed predictors.

Despite not being informed of the kinds of metrics that are useful for distinguishing nerve shapes, the discovered set of features seems to account for geometric features proposed in prior literature. Specifically, our PCs were mainly correlated to centerline length, curvature, cross-sectional area, and flattening ratio, similar to those that Ishiwada et al. found to be significantly different between the symptomatic and asymptomatic side on pre-operative imaging.[10] The convergence of these two sets of features provides a biological validation of the shape model learned by PCA. However, PCA is able to capture even more intricate features of the shape compared to conventional metrics, allowing the PCs to outperform conventional features in predictive models.

When independently analyzing the PCs that were most related to pain symptoms, the PCs associated with overall nerve size and flatness of the nerve were the most correlated with pain. Nerves on the side with TN pain were smaller and flatter than contralateral nerves. These findings are consistent with prior small-scale studies, which also commonly attribute the differences in shape to nerve atrophy related to the effects of neurovascular compression.[5,7,10,17] However, we also found that nerve size alone did not capture the full pain-associated morphology in our data, indicating that the higher-order shape features accounted for by our PCs provided additional information about symptoms. Furthermore, studies have also found that changes in nerve geometry after MVD may be related to favorable outcomes, suggesting shape may be a direct reflection of the processes causing nerve pain.[11]

Our study builds upon past studies in several ways. First, we validate these prior results in a large patient cohort. Additionally, the large cohort allows us to take a data-driven approach to discover important shape features. The analysis of a large dataset was critically enabled by our automated methods for nerve segmentation.[16] Data-driven methods can find disease-associated anatomical patterns that are not captured by simple measurements and have been widely applied to characterize disease across several medical fields.[3,4,15] In our study, a data-driven method found that pain was associated with multiple shape components, each of which correlated with multiple hand-crafted metrics. This insight suggests that the structural changes in TN may not be adequately captured by any single measurement and require higher-order shape features such as those found in our model.

A limitation of our method is that PCA is a linear decomposition method, finding the set of shape features that best fit the original nerve shapes when combined linearly. Some shape features, however, may be better captured by nonlinear methods. While PCA has the benefit of being more readily interpretable, more expressive decomposition techniques—such as variational autoencoders—may help capture these nonlinear shape features. Efforts to use more advanced methods, however, will need to balance the greater model flexibility with interpretability and generalizability.

Future work to assess these features specifically on patients who do not have apparent neurovascular conflict on pre-operative imaging will be important. Our current study only includes patients who underwent MVD, and as a result, nearly all patients had suspected neurovascular conflict pre-operatively. Since TN is a heterogeneous condition, analyzing an MVD cohort allows us to focus on a group of patients whose pain likely stems from a shared etiology. If the shape features found on this cohort are also found to be associated with pain in other cohorts, independent of neurovascular conflict, it would provide evidence that nerve shape is a biomarker of pain in TN more broadly even without offending vessels. Additionally, though we use a large cohort, our patients are from a single institution, and external validation will be important to evaluate the generalizability of our shape model.

## Conclusion

A data-driven approach to characterize trigeminal nerve morphology revealed multidimensional shape features that are associated with pain in trigeminal neuralgia. These shape features captured more pain-related anatomical information about the nerve compared to conventional geometric measures, suggesting that trigeminal nerve morphology may be an important biomarker of TN pain.

## Data Availability

Data and code used in the present study are available upon reasonable request to the authors.

## Conflict of Interest Statement

C.J. reported being a cofounder and advisor with equity interests for Egret Therapeutics and research funding from Biohaven, InCephalo, and Grifols. M.L. reported personal fees from Stryker during the conduct of the study. C.B. reported consulting for DePuy Synthes, Haystack Oncology, Privo Technologies, and Bionaut Labs and is a cofounder of OrisDx and Belay Diagnostics. No products from any of these companies were used in this study.

## Acknowledgements

This study did not receive any external funding sources. Generative artificial intelligence was used as a coding assistant in the data analysis process and was applied with human oversight and control. All outputs were reviewed, edited, and verified by the authors, who take full responsibility for the integrity and accuracy of the work. Data and code used in the present study are available upon reasonable request to the authors.

## Supplemental Figures

**Figure S1:**
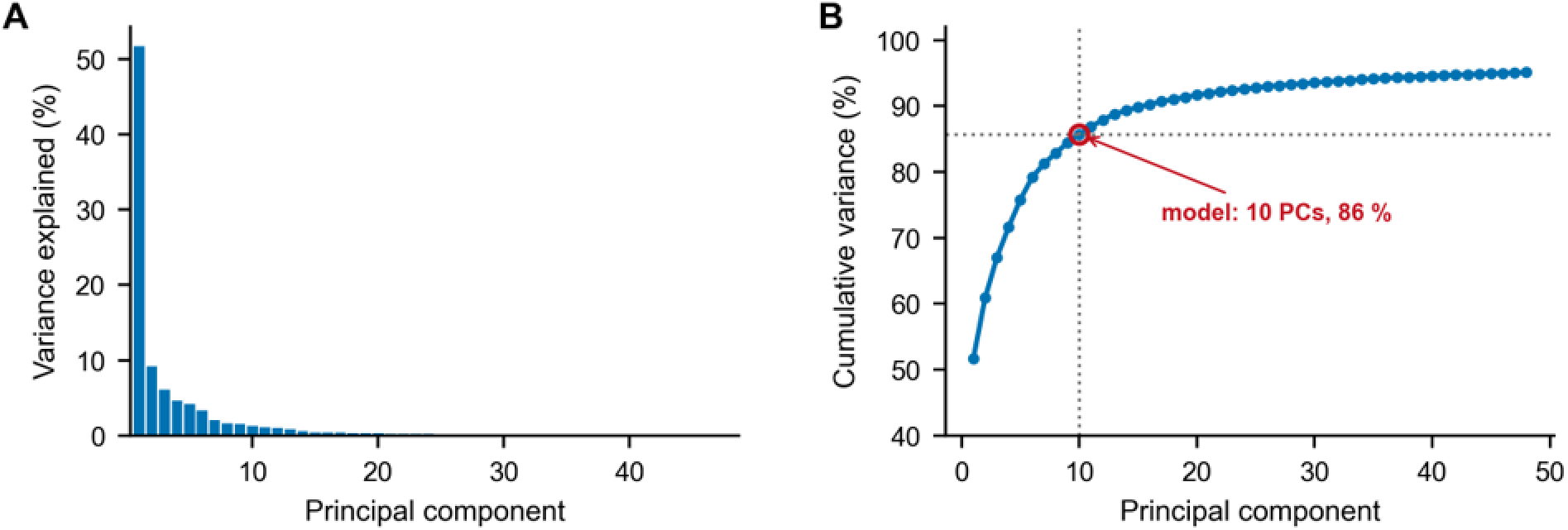
Variance explained by each principal component (A) and cumulatively (B) in the training set; the 10-component model explains 85.7% of the shape variance.

**Figure S2:**
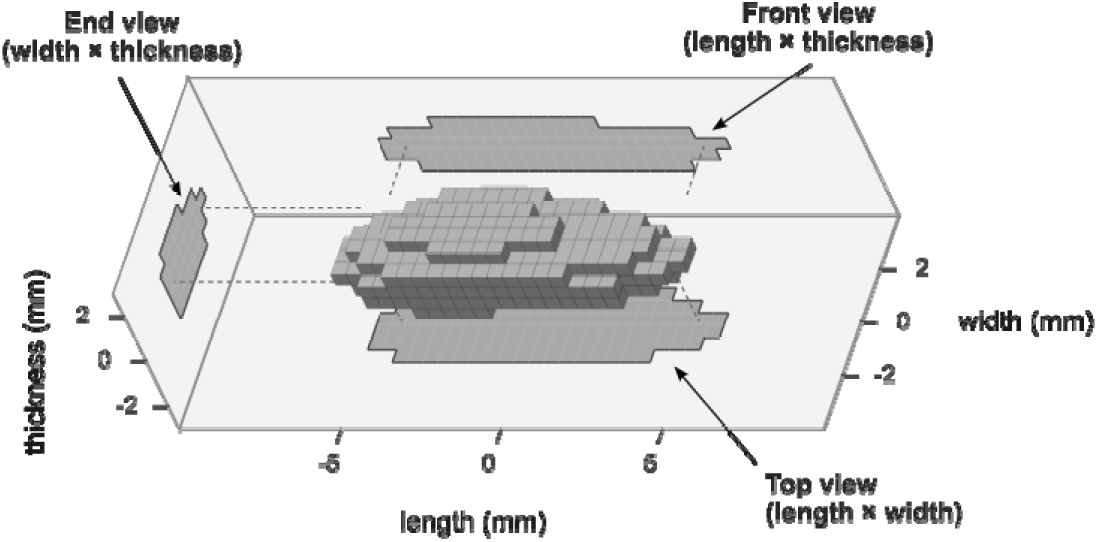
The mean of the training nerves is depicted, aligned along the three principal axes (length, width, and thickness). The three views of the nerve used in Figure 1 are shown.

**Figure S3:**
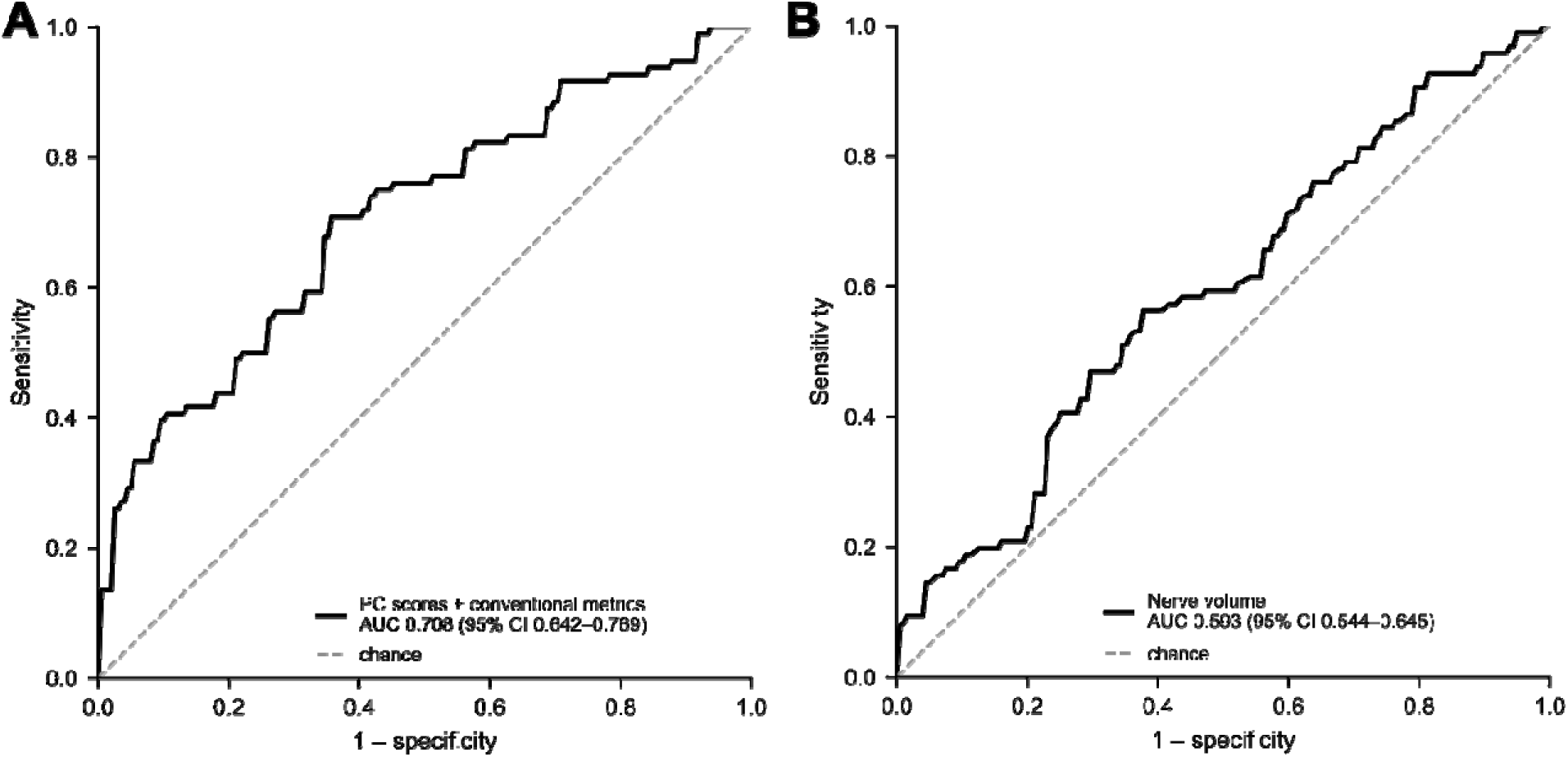
Receiver-operating characteristic (ROC) curves comparing logistic regression models for predicting whether a nerve is on the symptomatic side using the PC scores and conventional metrics together as predictors (A) and using total nerve volume as the only predictor (B).

## Notes

### Author Declarations

IRB of Johns Hopkins Medicine gave ethical approval for this work.

